# Spatial variation in excess mortality across Europe: a cross-sectional study of 561 regions in 21 countries

**DOI:** 10.1101/2023.04.04.23284990

**Authors:** Florian Bonnet, Pavel Grigoriev, Markus Sauerberg, Ina Alliger, Michael Mühlichen, Carlo-Giovanni Camarda

**Affiliations:** French Institute for Demographic Studies (INED), Aubervillers, France; Federal Institute for Population Research (BiB), Wiesbaden, Germany

## Abstract

**Objective:** To measure the burden of the COVID-19 pandemic in 2020 at the subnational level by estimating excess mortality, defined as the increase in all-cause mortality relative to an expected baseline mortality level.

**Design:** Statistical and demographic analyses of regional all-cause mortality data.

**Setting:** The vital statistics systems of 21 European countries.

**Participants:** The entire population of 561 spatial units in 21 European countries.

**Main Outcome Measures:** Losses of life expectancy at ages 0 and 60 for males and females.

**Results:** We found evidence of a loss in life expectancy in 391 regions, while only three regions exhibit notable gains in life expectancy in 2020. For 12 regions, losses of life expectancy amounted to more than 2 years, and three regions showed losses greater than 3 years. We highlight geographic clusters of high mortality in Northern Italy, Spain and Poland, while clusters of low mortality were found in Western France, Germany/Denmark and Norway/Sweden.

**Conclusions:** Regional differences of loss of life expectancy are impressive, ranging from a loss of more than 4 years to a gain of 8 months. These findings provide a strong rationale for regional analysis, as national estimates hide significant regional disparities.

**‘What is already known’:** Reported numbers of covid-19 deaths are subject to changes within and across countries due to inaccuracy, and incompleteness.

Excess mortality measured by loss in life expectancy is widely considered a relevant indicator for assessing the total mortality impact of the COVID-19 pandemic.

Whereas national estimates has been largely provided, few scattered regional studies for specific countries have been conducted.

**‘What this study adds’:** This study provides the first coherent analysis of excess mortality at regional level covering a large number of European countries.

It allows to properly map COVID-19 pandemic using official mortality data routinely collected by vital registration systems, which are less sensitive to misclassification.

**How this study might affect research, practice or policy’:** This study provides a strong rationale for regional analysis, as national estimates hide significant regional disparities

## 1. Introduction

The outbreak of SARS-CoV-2 triggered a strong reaction from public authorities, who responded in accordance with the threat that this new infectious disease entailed for the population. Despite having adopted social distancing measures that had never been seen in living memory in the 21st century, this pandemic led to most countries seeing significantly increased mortality [1], which the national surveillance authorities in Europe highlighted daily with their published reports [2, 3]. Nowadays, the daily monitoring of the pandemic’s evolution has been partially replaced by the quest to globally assess its mortality burden.

However, because these various national surveillance authorities were quickly established ad hoc to monitor the pandemic, use of the data they provide is broadly questioned for a wide range of reasons: different definitions of data among countries; time-varying collection methods; reporting delays; and diverse coverage by place of death [4, 5]. In contrast, the standard official statistics systems provide all-cause deaths, which are essential for computing excess mortality, which is defined as “the difference between the number of deaths (from any cause) that occur during the pandemic and the number of deaths that would have occurred in the absence of the pandemic” [6]. Excess mortality is considered by scholars to be the gold standard for estimating the global impact of SARS-CoV-2 [7, 8], which is why numerous studies have estimated excess mortality for a large number of countries [9–18].

While some of these studies evaluate the pandemic’s impact on mortality by estimating the absolute or relative numbers of excess deaths these figures are generally insufficient because they fail to consider changes in a population’s age structure. For this reason, scholars prefer looking at age-specific mortality rates instead, as they can be collapsed into a summary measure such as the age-standardised death rate or period life expectancy at birth (hereinafter, shortened to simply “life expectancy”). Especially, the latter measure is a popular tool for quantifying period shocks in mortality because life expectancy readily expresses changes in mortality in terms of a longer or shorter average life span and are thus, comprehensible for a broad audience. The impact on changes in age-specific mortality rates on life expectancy trends is, however, more complex than often assumed (see e.g., [19–21]). Recently, [22] suggested interpreting a drop in life expectancy due to a mortality shock such as those incurred by the COVID-19 pandemic rather as a measure of premature mortality as compared to differences in mean longevity. This new interpretation might be suitable for our analysis because the mortality conditions observed in 2020 at the subnational level are unlikely to prevail in the future and thus, it is difficult to conceive the presented mortality changes as differences in average life spans [23].

To date, most of these studies estimate excess mortality for countries as a whole, with the undeniable risk of hiding intranational differences and hindering effective health policies within a pandemic context. Hence, in recent months, many papers have estimated regional excess mortality for specific countries [24–32]. However, comparing these regional patterns is relatively problematic because these studies take different approaches to compute the mortality levels that would have occurred without the pandemic. Specifically, they either use pre-pandemic levels or employ forecasting techniques. Moreover, these papers rely on different indicators to assess excess mortality, namely life expectancy or death toll. To our knowledge, only one study [33] allows for a simultaneous comparison of regional excess deaths in five European countries (Italy, Switzerland, Spain, Greece, England, and Wales) in 2020.

Our paper aims to fill this research gap by presenting life expectancy losses at birth in 2020 for 561 regions from 21 countries in Central and Western Europe. In dealing with small geographical areas, we focus on point-estimates as much as on uncertainty quantification. In order to obtain the mortality levels that would have been observed in 2020 in the absence of the pandemic, we took a robust forecasting approach that accounts for regional diversity and delivers analytic confidence intervals that account for all sources of uncertainty [34, 35]. Furthermore, this study also identifies geographical clusters of high and low losses of life expectancy.

## 2. Data and Methods

### 2.1. Data Preparation

We collected regional death and population counts for 21 European countries by age and sex from the national statistical offices, Eurostat, and the Human Mortality Database [36]. Since these data show varying age classes, we harmonised them into single-age intervals up to 95+ for all spatial units [37]. The lowest number of age groups in our input data is sixteen (for Slovakia) and the largest age group that we ungrouped into single ages is fourteen (for Germany, deaths at age 1 to 14). To ensure consistency in area size when selecting spatial units among the 21 countries, we relied on mostly Nomenclature of Territorial Units for Statistics (NUTS) 3 (Czechia, Denmark, France, Italy, Luxembourg, Slovakia, Spain, Sweden) and NUTS 2 levels (Austria, Belgium, Hungary, Iceland, Netherlands, Norway, Portugal, Switzerland, Slovenia, England, Wales). As exceptions, we used NUTS 1 for Ireland, Northern Ireland, and Scotland; and, for Germany, a national classification (“Raumordnungsregionen”). Minor adjustments were required due to territorial changes over time and data availability issues (see online supplementary Appendix A, Table 1 for details). In total, we analysed 561 harmonised spatial units containing populations ranging from 40,000 (Bornholm, Denmark) to 6,760,000 (Madrid, Spain).

Finally, we verified the data quality by comparing our regional data (aggregated at the national level) with those available in the Human Mortality Database. The differences between age-specific death rates calculated from the two data sources are almost nil for ages below 90, and very small for ages upper this limit. Thus, summary measures such as life expectancy at birth correspond to each other.

### 2.2 Methodology

When dealing with excess mortality, a central methodological issue concerns estimating the baseline mortality level, which is what would have been the expected level in the absence of the pandemic. Pre-pandemic mortality levels are often used as the baseline because they are easy to obtain and compute. However, because such a simplistic approach frequently ignores temporal trends, it is necessary to derive a more suitable expected mortality level in the absence of COVID-19 by forecasting pre-pandemic historical trends for the year affected by the pandemic (here, 2020). Among the numerous methodologies currently available (e.g., [38]), we opted for a CP-spline approach [35], which combines two-dimensional P-splines with prior demographic information derived from population-specific historical patterns.

A significant advantage to taking a non-parametric approach like that of CP-splines lies in its great flexibility for describing various mortality scenarios, which becomes particularly relevant when dealing with 561 diverse subpopulations across 21 European countries. Furthermore, smooth, plausible age profiles and time trends are ensured while we gain the added advantage of robustness when analysing small populations at risk. Moreover, the relatively low computation costs of CP-splines allow us to optimise region-specific time windows for our 2020 forecast values.

It is essential to quantify uncertainty before drawing any conclusions on excess mortality levels, particularly when dealing with regional mortality data. The error associated with the forecasting procedure must be added to the uncertainty around the observed 2020 mortality levels. Whereas the variance-covariance structure from CP-splines allows us to compute confidence intervals for our forecast values, a fully analytic procedure is used to compute uncertainty around the observed 2020 mortality levels. These two sources of uncertainty are then combined without performing any simulation or bootstrap procedures, thereby considerably reducing computational costs and time. A detailed description of the analytic procedure is available in online supplementary Appendix C.

It is noteworthy that the whole procedure can be performed for any given age and regardless of the mortality indicator selected for estimating excess mortality (e.g., life expectancy or age-standardised death rate). More details on this approach can be found in [34]. All calculations are performed in R version 4.0.3 [39].

To identify any spatial clustering of losses of life expectancy, we rely on the Getis-Ord Gi* statistic [40], which allows us to locate hot and cold spots, that is, neighbouring areas of high concentration of spatial units with either large or small loss of life expectancy. In practice, Gi* is calculated for all estimated losses of life expectancy in our dataset, and the resultant z-scores and p-values are evaluated within a defined neighbourhood. Here, we used the contiguity edges corners definition of neighbourhood, which specifically considers neighbours to be units that share a boundary, share a node, or overlap. To be a statistically significant hot spot, a spatial unit should not only have a high value but also be surrounded by other units with high values. The local sum for a given unit and its neighbours is evaluated against the sum of all spatial units. If the observed local sum is very different from the expected local sum, then this difference is not the result of random chance, and the corresponding z-score is statistically significant. The same logic is applied for identifying cold spots. This well-established methodology has been implemented and thoroughly documented in ArcMap software (module *Spatial Statistics Tools*) [41], which we used to perform the respective analysis as well as to produce mortality maps. Required shapefiles were collected from Eurostat [42] and were re-projected using the *Winkel tripel* projection.

## 3. Results

Figure 1 shows the spatial distribution of losses of life expectancy at birth across 561 territorial units in 21 European countries during 2020. Light blue represents gains in life expectancy compared to expected values, whereas the remaining colours indicate life expectancy losses. Among all the countries considered in our analysis, those most affected by the pandemic appear to be Italy, Spain, and Poland. In most Polish regions as well as in many Spanish and Italian regions, losses in both male and female life expectancy amounted to more than 1 year. This fall in life expectancy observed within just one calendar year is unprecedented in 21^st^ century Europe [9, 10]. Interestingly, they are among the areas with both the highest longevity (Spanish and Italian regions) and the lowest longevity (Polish regions) in Europe (see online supplementary Appendix A, Figures A1 and A2). In contrast, most regions in Denmark, Germany, Norway, and western France, as well as some rural areas in Sweden, showed only moderate declines or even slight increases in life expectancy in 2020 compared to their expected values.

**Figure 1.**
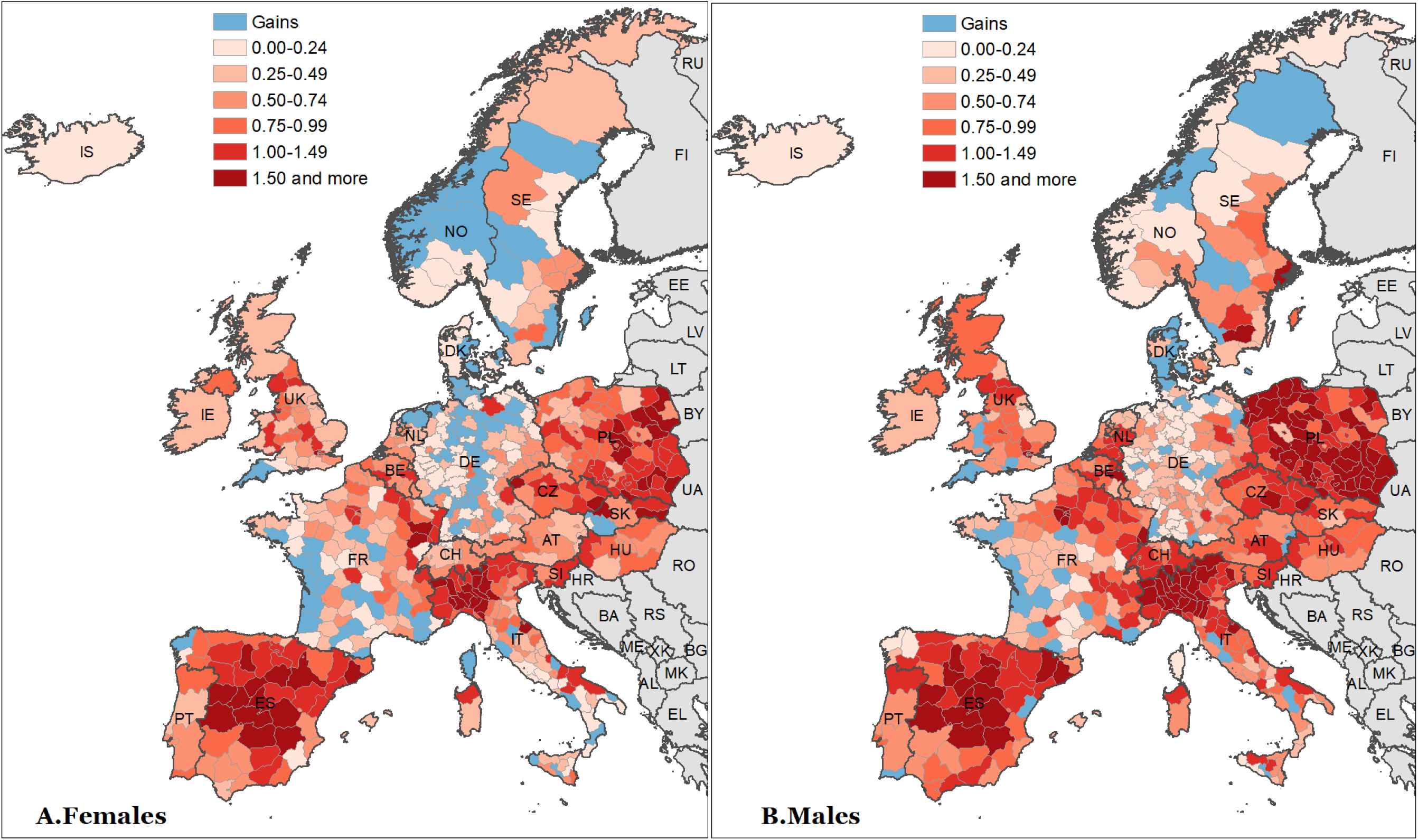
Spatial distribution of losses of life expectancy at birth (years) across 21 European countries in 2020 Source: As shown in Table A1.

**Figure 2.**
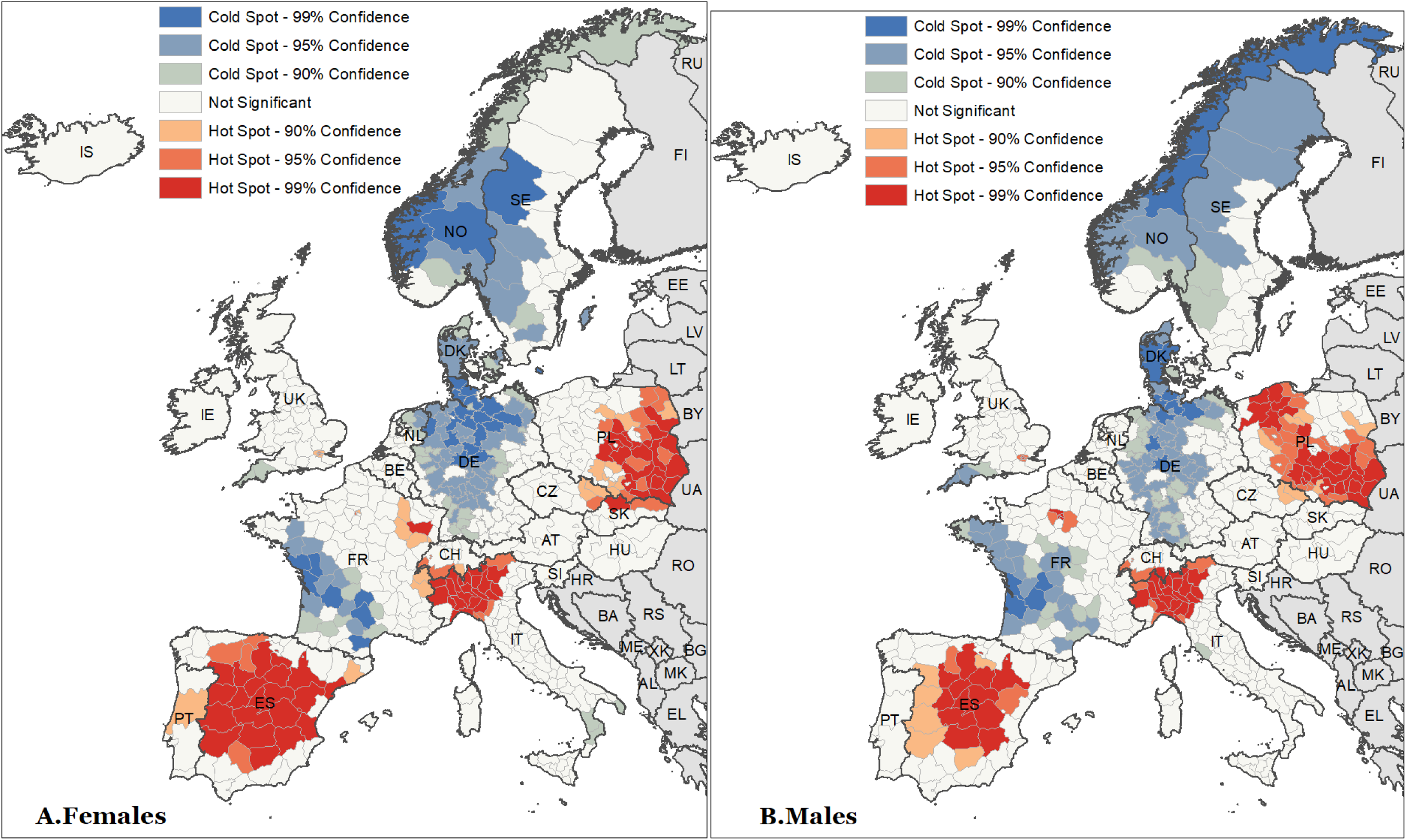
Hot and cold spots of losses of life expectancy at birth across 21 European countries in 2020. Source: As shown in Table A1.

Although previous works could only show national differences in excess mortality associated with the COVID-19 pandemic, our regional analysis is able to highlight substantial within-country variations at the subnational level. Italy represents a notable example, with the whole country showing a loss of life expectancy at birth of 1.16 years for both sexes combined. However, this national value conceals both a loss of 4.18 years in the Bergamo province in the north (CI: −3.95; −4.4) and a gain of 0.4 years (CI: −0.17; 0.92) in the Sicilian province of Caltanissetta. Moreover, we can see in Figure 3 that 95% confidence intervals are plotted along estimated excess mortality in life expectancy at birth. Other examples of remarkable subnational differences can be found in France and the United Kingdom. In the former case, higher excess mortality is visible in the north-east compared to the south-west regions. In the latter, south-west shows a moderate gain, unlike the central parts of England that experience high losses. Spatial variation is also evident in Germany, a country with relatively better performance during the first pandemic wave in 2020. Specifically, its eastern and south-eastern regions bordering Poland, Czechia, and Austria show higher life expectancy losses than the rest of the country.

**Figure 3.**
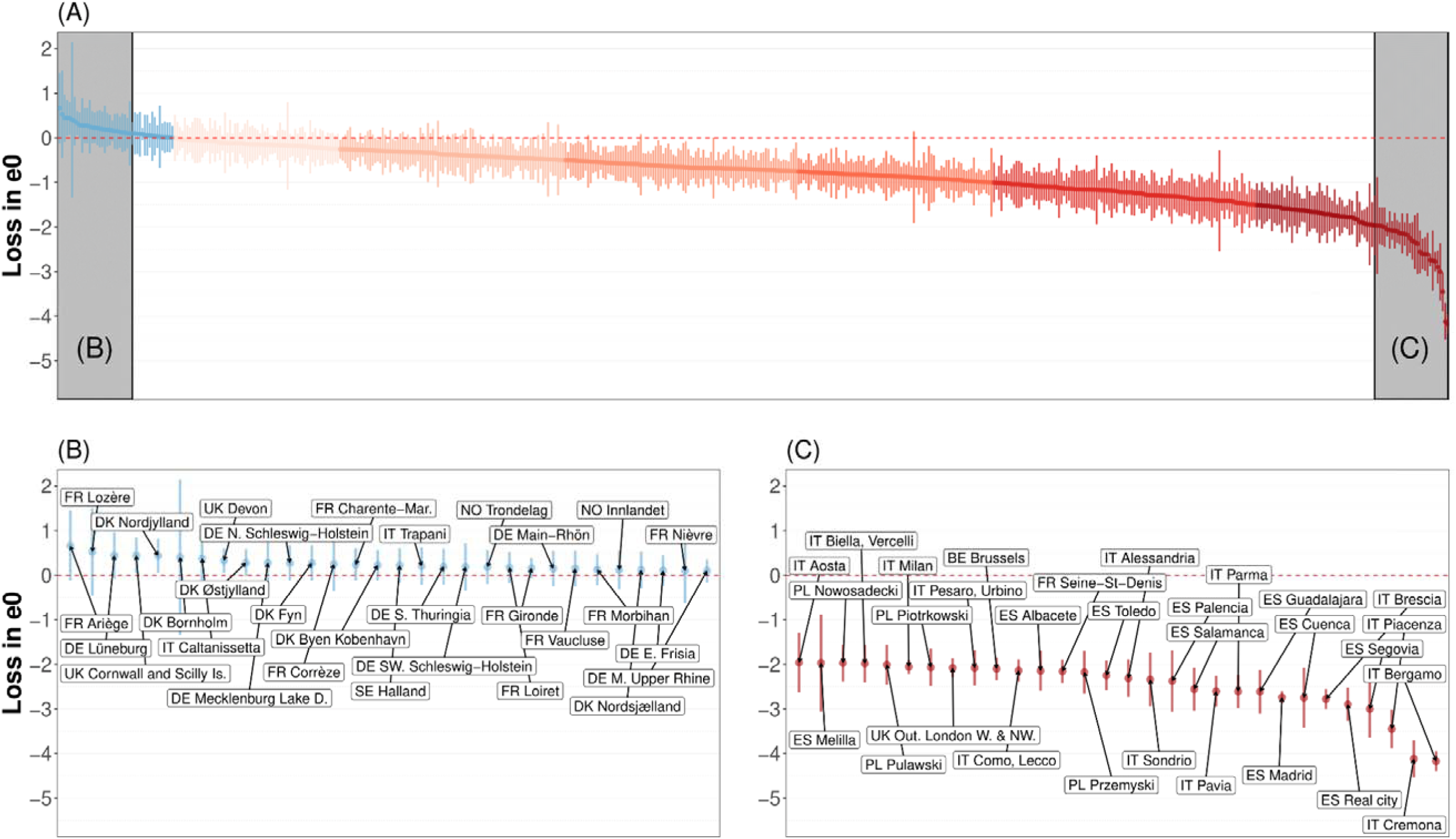
Losses or gains of life expectancy at birth (e0) associated with COVID-19 pandemic across 561 spatial units in 21 European countries, 2020, both sexes combined. Source: As shown in Table A1. Notes: 1) Excess mortality is estimated as the difference between observed and expected life expectancy at birth 2) Vertical bars represent 95% confidence intervals.

In contrast, although Poland shows life expectancy losses similar to Italy (1.32 year), there is considerably less subnational variation between the highest and lowest affected regions, with Jeleniogorski in the south showing a loss of 0.7 years (CI: −0.16; −1.09) and Przemyski in the east a loss of 2.17 years (CI: −1.7; −2.65).

The analysis of spatial distribution of losses of life expectancy at age 60 has revealed very similar results (online supplementary Appendix A, Figure A3).

Our next step was to analyse the hot and cold spots for losses of life expectancy at birth (Figure 2) and at age 60 (online supplementary Appendix A, Figure A4). This analysis statistically confirms our previous visual inspection: Hot spots of life expectancy losses were indeed located in northern Italy and covered most of the Polish and Spanish territory; whereas cold spots were located in Denmark, Germany, western France, and Norway. Furthermore, our analysis identifies several small clusters of elevated excess mortality expanding across national borders. One example can be seen in an area covering parts of northern Italy and southern Switzerland, and – at least among women – spreading into eastern France. Another cross-national hot spot expands from southern Poland into eastern Czechia and – again, among women – into northern Slovakia. These observations may well indicate possible spill-over effects of the pandemic burden across European countries.

Figure 3 ranks the life expectancy losses and shows the associated 95% confidence intervals (CI) for all 561 spatial units. This figure also allows us to visually assess the magnitude of uncertainty around point estimates. The Panel A colours correspond to the colours and cut points presented in Figure 1. Panels B and C depict the top and bottom 30 regions, respectively. Figures A5 and A6 in the online supplementary Appendix A separately show the same distributions for men and women.

Figure 3 also reveals the large regional variability in excess mortality across Europe. The aforementioned Italian Bergamo province experienced the highest loss of life expectancy, amounting to 4.18 years (CI: −3.95; −4.4); whereas the Ariège department in France benefited from the highest gain of 0.65 years (CI: −0.13; 1.45). Only a few regions experienced life expectancy gains for which the 95% confidence intervals do not include 0: Devon, Cornwall, and Scilly Island in the UK; and Nordjylland in Denmark. Overall, 391 out of 561 regions suffered consequential life expectancy losses, meaning that the associated 95% confidence intervals do not contain 0. The tail of the distribution (Panel C) is dominated by northern Italian and central Spanish regions. Among those harshly hit by the pandemic, 12 regions experienced losses of life expectancy greater than 2 years, with the Italian provinces of Piacenza, Cremona, and Bergamo experiencing more than 3 years of losses. As expected, the group of regions least affected by the pandemic (Panel B) is dominated by German, French, and Danish spatial units. However, this successful group also includes some Italian and UK regions. The numerical values of losses of life expectancy at birth and at age 60 for each spatial unit and for both females and males are provided in the online supplementary Appendix B.

In addition to revealing intranational heterogeneity and cross-national patterns, using subnational data has the advantage of enabling a deeper analysis of excess mortality. Moreover, the uncommonly large number of spatial units also increases statistical power in these supplementary analyses. For instance, we can easily assess the association between sex differences in life expectancy loss and the overall regional loss. Figure 4 displays the results of this closer examination using a modified version of a Bland-Altman plot, where the differences between female and male changes in life expectancy are plotted against the corresponding overall regional change. Each dot in the plot is sized proportionally to the population of its respective region.

**Figure 4.**
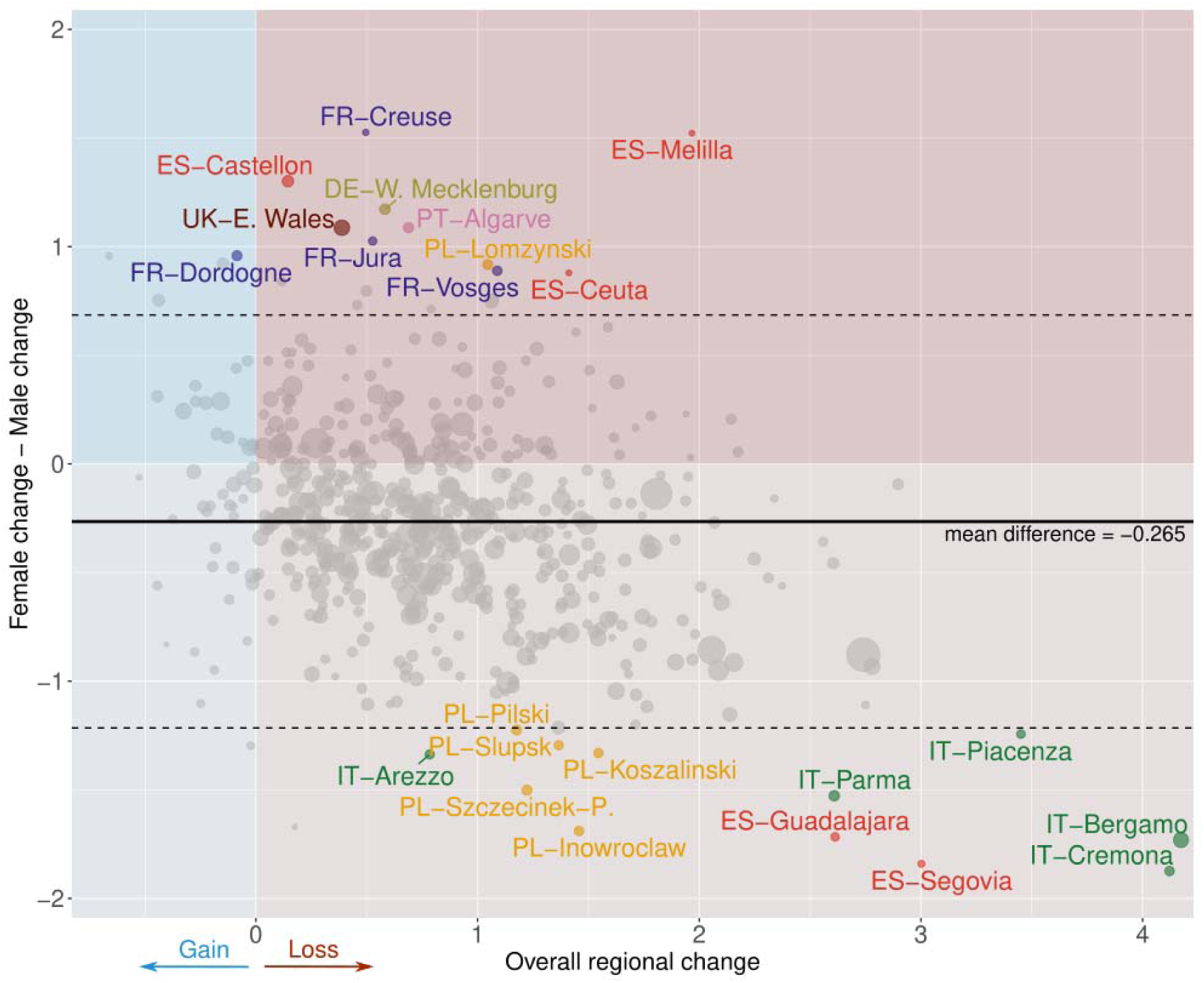
A modified version of a Bland-Altman plot to portray association between differences between female and male losses of life expectancy at birth and overall regional changes across 561 spatial units in 21 European countries, 2020. Source: As shown in Table A1.

As already clear from previous analysis, the majority of regions present an overall loss in life expectancy and this is indicated by the large number of dots in the red areas of Figure 4. The average difference in excess mortality between males and females is clearly indicated by the mean line, showing that males have experienced over three months of additional loss on average. Regions with higher losses among males are represented by dots below the horizontal zero line (lighter red), while regions with greater losses among females are indicated by dots above this line (darker red).

The identification of outliers is particularly noteworthy. Here, we identify outliers by depicting spatial units lying beyond the 95% limits of agreement of the Bland-Altman plot estimated as prediction intervals of a mean model. Notably, nearly all outliers among regions with greater losses of males belong to areas identified as hot spots (Italy, Poland, Spain). Nevertheless, some outliers also show greater female losses, and these are located predominantly in France.

## 4. Discussion

### 4.1. Statement of Principal Findings

This large study of 561 territorial units in 21 countries uses loss of life expectancy to measure the burden of COVID-19 in 2020 throughout Western and Central Europe. Our findings show evidence of losses in 391 regions, while only three regions exhibit notable gains in life expectancy during the first year of pandemic. Overall, those most affected were central Spain, northern Italy, and eastern Poland, while Denmark, western France, Germany, and Norway were somewhat spared from the COVID-19 burden. Regional differences in life expectancy changes were impressive, ranging from a loss of more than 4 years to a gain of 8 months. For 12 regions, losses of life expectancy amounted to more than 2 years, and three regions showed losses greater than 3 years.

This study further reveals that regional differences in excess mortality were also highly substantial within some countries. In Italy, for example, we estimated a 4-year life expectancy loss in the Bergamo province but no loss in the Caltanissetta province. However, the regional differences in one country can differ greatly from those in another. For example, although Italy and Poland both have similar losses of life expectancy at birth at the national level, the gap between the highest and lowest afflicted regions is three times larger in Italy than in Poland.

Geographic clusters identify where life expectancy was much lower than expected (hot spot), and others where life expectancy was in line with expectations or even higher (cold spot). Notwithstanding, these analyses pinpoint clusters where excess mortality expanded beyond national borders in certain European areas: The hot spots observed in northern Italy spilled over into southern Switzerland and eastern France, and other hot spots observed in Poland spilled over into northern Slovakia and eastern Czechia. Meanwhile, this study singles out a cold spot located in two nearby countries (Germany and Denmark).

We observe differences between women and men in their life expectancy losses in 2020. In most analysed regions, reduced life expectancy was greater among men. For example, in the worst hit regions in northern Italy and central Spain, men’s losses were more than 1 year higher as compared to their female counterparts. Widening sex differentials in mortality during the COVID-19 pandemic have been reported previously [43, 44, 1]. Yet, we also find more pronounced life expectancy losses for women in many European regions where drops in life expectancy were more moderate. This is relevant for the discussion on causes for male excess deaths, i.e., biological mechanisms vs. social mechanism [45]. As we observe substantial variability in the magnitude and even direction of sex differences in life expectancy losses, our results indicate that the degree of mortality deterioration is not necessarily linked to biological sex. Further, it complements the findings by [1] who also identified higher life expectancy losses in several European countries but failed to find statistically significant changes because they only looked at the national level.

### 4.2. Strengths and Limitations

This study is the first to use a large set of small territorial units to provide estimates of loss of life expectancy during the COVID-19 pandemic in 2020. The contiguity of the regions investigated has allowed us to accurately identify geographic clusters of elevated mortality. This approach is novel because most previous studies have focused primarily on single-country regions or on regions in various countries with no common borders.

Contrary to many other studies on COVID-19-related deaths, we estimated losses of life expectancy using official mortality data routinely collected by vital registration systems, which are less sensitive to reporting delays and misclassification. To verify that each country’s sums of its regional data were consistent for all ages, we verified them with data from the Human Mortality Database. This is especially crucial, not only for calculating outcomes at older ages, which require statistical techniques to obtain single-year-of-age data, but also considering how COVID-19 was particularly dangerous for older individuals. Finally, we computed baseline mortality using an up-to-date statistical methodology that optimises the time windows in our models for forecasting regional trends in 2020. We compared our country-specific estimates with results from previous studies in order to evaluate our findings in terms of reliability. Our estimates for losses in life expectancy are consistent with results from Aburto et al. [9] and Islam et al. [15] (see table A2).

Our study also has several limitations. It covers 21 Central and Western European countries and still cannot be extended to Eastern European countries. However, comparisons with these countries would have been interesting insofar as this global health crisis triggered different political responses. Furthermore, this study covers only the year 2020. Given the experience gained by policymakers during the first waves of the pandemic and the emergence of new variants, the results for 2021 and 2022 should be different. As soon as more up-to-date data become available, the analyses we performed can be easily extended along both the spatial and temporal dimensions.

While our adopted forecast approach is robust and flexible enough to be adapted to different demographic scenarios and relatively small populations, we model each spatial unit independently and without considering spatial autocorrelation. By including a spatial structure in modelling and forecasting mortality, this may eventually reduce uncertainty around excess mortality estimates and thus lead to more explicit outcomes. In order to avoid underestimating excess mortality, it is paramount to account for temporal trends when estimating baseline mortality. However, other approaches in computing baseline mortality levels have been proposed [46–48], and these could be adopted in order to obtain alternative perspectives.

### 4.3. Policy Implications and Future Research

Our study extends the existing literature on excess mortality during the COVID-19 pandemic in 2020 by reporting results at the regional level for many European countries, which has not been done so far. The findings provide a strong rationale for this regional analysis, as we show that national estimates would have hidden significant regional disparities. Policymakers should be made aware of this intranational heterogeneity in order to fully assess the burden of the pandemic in their own countries and adopt differentiated health policy responses.

While our study quantifies regional differences in excess mortality, it cannot provide avenues for explaining them. The first step toward doing so would be to link these estimates to regional contextual variables (such as income level or occupational structure) as well as to public policies of social distancing and international isolation, which were implemented at both regional and national levels. Eventually, ecological analyses should be complemented with carefully designed epidemiological studies. In this way, one will be able to highlight the decisive factors explaining these regional differences and ultimately gain a better understanding of how to deal with the spread of a new infectious disease. For example, the exceptional mortality observed in northern Italy might be due to the early onset of the pandemic, which triggered a strong public response from the Italian government that limited the spread of the pandemic to the southern parts of the country [49].

It should be emphasised that this study estimates the burden of the pandemic by calculating excess mortality, which is the overall net assessment of the pandemic’s impact on society and must be distinguished from the mortality count due to COVID-19. More specifically, the Institute for Health Metrics and Evaluation (IHME) considers excess mortality to be influenced by six drivers related to the pandemic and the response to it, which are namely: (i) deaths directly related to SARS-CoV-2 infection; (ii) increased mortality due to cancelling or postponing care; (iii) increased mortality due to mental health problems, including depression, increased alcohol use, and increased opiate use; (iv) reduced mortality from decreases in accidents because of reduced mobility; (v) reduced mortality due to reduced transmission of other viruses, including influenza; and (vi) reduced mortality from many chronic diseases such as cardiovascular and respiratory diseases that would have killed frail individuals had COVID-19 not killed them instead. When detailed cause-of-death statistics become available, an accurate assessment must be made of COVID-19 mortality, as well as the distinction between direct and indirect COVID-19 mortality.

Finally, this study critically depends on reliable data being made available from the national statistical institutes of the different European countries analysed here. The magnitude of our presented results should encourage policymakers to promptly provide regional data in order to better measure the burden of future epidemics in a timely fashion. In order to produce accurate analyses in times of demographic ageing, it is of utmost importance to harmonise European population and mortality data with consistent age groups and an oldest age class of at least 100 years upwards.

## Data Availability

All data produced are available online at :

https://osf.io/h68wz/?view_only=47353f6f4b2e41cab3c761246e59d615

## Acknowledgments

We are very grateful to Markéta Majerová, Rok Hrzic, Magdalena Muszyńska-Spielauer, and Mathias Lerch for providing, respectively, Czech, Slovenian, Austrian, and Swiss data.

## Online Supplementary Appendix A

**Table A1:** Regional division, sources, data information and adjustments by country.

| Country | Spatial units | Source | Period | Upper age limit | Adjustments/comments |
| --- | --- | --- | --- | --- | --- |
| Austria | 9 NUTS 2 units ('Länder') | Statistics Austria | 1990–2020 | Deaths: 90<br>Pop.: 100 | – |
| Belgium | 11 NUTS 2 units ('provinces') | Belgian Statistical Office | 1992–2020 | Deaths: 100<br>Pop.: 100 | – |
| Czechia | 14 NUTS 3 units ('kraje') | Czech Statistical Office | 1996–2020 | Deaths: 95<br>Pop.: 95 | – |
| Denmark | 11 NUTS 3 units ('landsdele') | Statistics Denmark | 2008–2020 | Deaths: 100<br>Pop.: 100 | – |
| France | 95 NUTS 3 units ('départements') | INSEE | 1970–2020 | Deaths: 105<br>Pop.: 105 (after 2016: 95) | Non-European areas excluded; north and south of Corse merged to maintain consistent time series |
| Germany | 96 ROR ('Raumordnungs-regionen') | Statistical Offices of the German Länder | 1992–2020 | Deaths: 90<br>Pop.: 90 | Harmonised to apply current territorial administrative division (as of Dec 2022) to the whole study period and to eliminate the Census 2011 break; 400 NUTS 3 units ('Kreise') aggregated to 96 ROR units according to classification of BBSR (2017) |
| Hungary | 8 NUTS 2 units ('tervezési-statisztikai régiók') | Hungarian Central Statistical Office, Eurostat | 2001–2020 | Deaths: 90<br>Pop.: 90 | – |
| Iceland | 1 NUTS 2 unit | Human Mortality Database | 1970–2020 | Deaths: 110<br>Pop.: 110 | – |
| Ireland | 1 NUTS 1 unit | Human Mortality Database | 1990–2020 | Deaths: 110<br>Pop.: 110 | No subnational division due to data availability issues |
| Italy | 92 NUTS 3 units ('province') | ISTAT | 1995–2020 | Deaths: 100<br>Pop.: 100 | We merged the following regions to maintain a consistent time series: 1) Biella + Vercelli, 2) Novara + Verbano, 3) Como + Lecco, 4) Milano + Lodi + Monza + Brianza, 5) Rimini + Forli-Cesena, 5) Firenze + Prato, 6) Cagliari + Medio Campidano + Carbonia-Iglesias + Ogliastra + Oristano + Nuoro, 7) Sassari + Olbia-Tempio, 8) Foggia + Bari + Barletta, 9) Fermo + Ascoli-Piceno, 10) Crotone + Vibo Valentia + Cantanzaro. |
| Luxembourg | 1 NUTS 3 unit | Human Mortality Database | 1996–2020 | Deaths: 110<br>Pop.: 110 | – |
| Netherlands | 11 NUTS 2 units ('provinces') | Statistics Netherlands | 2001–2020 | Deaths: 100<br>Pop.: 100 | – |
| Norway | 7 NUTS 2 units ('landsteder') | Statistics Norway | 2000–2020 | Deaths: 100<br>Pop.: 105 | Harmonised to apply current territorial administrative divisions; we excluded remote islands Svalbard and Jan Mayen |
| Poland | 73 NUTS 3 units ('podregiony') | Statistics Poland | 2006–2020 | Deaths: 85<br>Pop.: 85 (until 2005: 65) | – |
| Portugal | 5 NUTS 2 units ('regiões') | National Institute of Statistics, Eurostat | 1992–2020 | Deaths: 85<br>Pop.: 95 | For visibility reasons, we excluded the remote islands Azores and Madeira. |
| Slovakia | 8 NUTS 3 units ('kraje') | Slovakian Statistical Office | 1996–2020 | Deaths: 85<br>Pop.: 110 | – |
| Slovenia | 2 NUTS 2 units ('kohezijne') | Slovenian Statistical Office | 2002–2020 | Deaths: 100<br>Pop.: 100 | – |
|  | regije') |  |  |  |  |
| Spain | 50 NUTS 3 units ('provincias') | National Statistics Institute | 1990–2020 | Deaths: 100<br>Pop.: 100 (until 2001:85) | For visibility reasons, we excluded the Canary Islands. |
| Sweden | 21 NUTS 3 units ('län') | Statistics Sweden | 1969–2020 | Deaths: 100<br>Pop.: 100 | – |
| Switzerland | 7 NUTS 2 units ('Grossregionen') | Federal Statistical Office | 1991–2020 | Deaths: 95<br>Pop.: 100 | – |
| United Kingdom | 37 NUTS 2 regions | Office for National Statistics (for England & Wales); Human Mortality Database (for Northern Ireland & Scotland) | 2002–2020 | England & Wales<br>Deaths: 90<br>Pop.: 90<br>Northern Ireland & Scotland<br>Deaths: 110<br>Pop.: 110 | NUTS 1 level data for Northern Ireland and Scotland due to data quality issues |

**Table A2:**
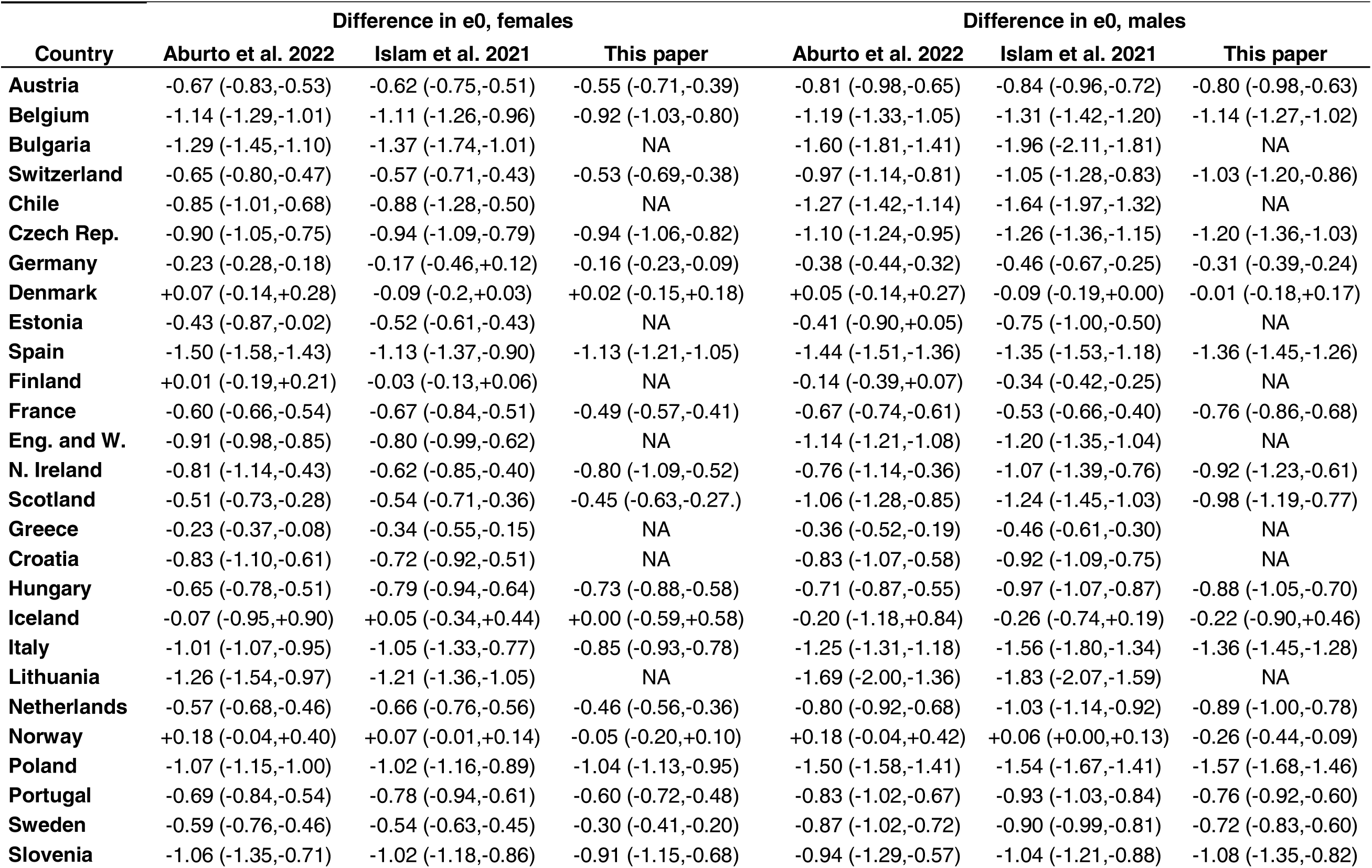
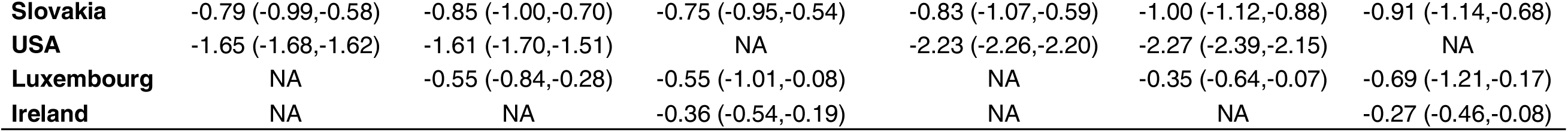
Comparison of estimated life expectancy changes with previous studies.

| Country | Difference in e0, females |  |  | Difference in e0, males |  |  |
| --- | --- | --- | --- | --- | --- | --- |
|  | Aburto et al. 2022 | Islam et al. 2021 | This paper | Aburto et al. 2022 | Islam et al. 2021 | This paper |
| **Austria** | -0.67 (-0.83,-0.53) | -0.62 (-0.75,-0.51) | -0.55 (-0.71,-0.39) | -0.81 (-0.98,-0.65) | -0.84 (-0.96,-0.72) | -0.80 (-0.98,-0.63) |
| **Belgium** | -1.14 (-1.29,-1.01) | -1.11 (-1.26,-0.96) | -0.92 (-1.03,-0.80) | -1.19 (-1.33,-1.05) | -1.31 (-1.42,-1.20) | -1.14 (-1.27,-1.02) |
| **Bulgaria** | -1.29 (-1.45,-1.10) | -1.37 (-1.74,-1.01) | NA | -1.60 (-1.81,-1.41) | -1.96 (-2.11,-1.81) | NA |
| **Switzerland** | -0.65 (-0.80,-0.47) | -0.57 (-0.71,-0.43) | -0.53 (-0.69,-0.38) | -0.97 (-1.14,-0.81) | -1.05 (-1.28,-0.83) | -1.03 (-1.20,-0.86) |
| **Chile** | -0.85 (-1.01,-0.68) | -0.88 (-1.28,-0.50) | NA | -1.27 (-1.42,-1.14) | -1.64 (-1.97,-1.32) | NA |
| **Czech Rep.** | -0.90 (-1.05,-0.75) | -0.94 (-1.09,-0.79) | -0.94 (-1.06,-0.82) | -1.10 (-1.24,-0.95) | -1.26 (-1.36,-1.15) | -1.20 (-1.36,-1.03) |
| **Germany** | -0.23 (-0.28,-0.18) | -0.17 (-0.46,+0.12) | -0.16 (-0.23,-0.09) | -0.38 (-0.44,-0.32) | -0.46 (-0.67,-0.25) | -0.31 (-0.39,-0.24) |
| **Denmark** | +0.07 (-0.14,+0.28) | -0.09 (-0.2,+0.03) | +0.02 (-0.15,+0.18) | +0.05 (-0.14,+0.27) | -0.09 (-0.19,+0.00) | -0.01 (-0.18,+0.17) |
| **Estonia** | -0.43 (-0.87,-0.02) | -0.52 (-0.61,-0.43) | NA | -0.41 (-0.90,+0.05) | -0.75 (-1.00,-0.50) | NA |
| **Spain** | -1.50 (-1.58,-1.43) | -1.13 (-1.37,-0.90) | -1.13 (-1.21,-1.05) | -1.44 (-1.51,-1.36) | -1.35 (-1.53,-1.18) | -1.36 (-1.45,-1.26) |
| **Finland** | +0.01 (-0.19,+0.21) | -0.03 (-0.13,+0.06) | NA | -0.14 (-0.39,+0.07) | -0.34 (-0.42,-0.25) | NA |
| **France** | -0.60 (-0.66,-0.54) | -0.67 (-0.84,-0.51) | -0.49 (-0.57,-0.41) | -0.67 (-0.74,-0.61) | -0.53 (-0.66,-0.40) | -0.76 (-0.86,-0.68) |
| **Eng. and W.** | -0.91 (-0.98,-0.85) | -0.80 (-0.99,-0.62) | NA | -1.14 (-1.21,-1.08) | -1.20 (-1.35,-1.04) | NA |
| **N. Ireland** | -0.81 (-1.14,-0.43) | -0.62 (-0.85,-0.40) | -0.80 (-1.09,-0.52) | -0.76 (-1.14,-0.36) | -1.07 (-1.39,-0.76) | -0.92 (-1.23,-0.61) |
| **Scotland** | -0.51 (-0.73,-0.28) | -0.54 (-0.71,-0.36) | -0.45 (-0.63,-0.27.) | -1.06 (-1.28,-0.85) | -1.24 (-1.45,-1.03) | -0.98 (-1.19,-0.77) |
| **Greece** | -0.23 (-0.37,-0.08) | -0.34 (-0.55,-0.15) | NA | -0.36 (-0.52,-0.19) | -0.46 (-0.61,-0.30) | NA |
| **Croatia** | -0.83 (-1.10,-0.61) | -0.72 (-0.92,-0.51) | NA | -0.83 (-1.07,-0.58) | -0.92 (-1.09,-0.75) | NA |
| **Hungary** | -0.65 (-0.78,-0.51) | -0.79 (-0.94,-0.64) | -0.73 (-0.88,-0.58) | -0.71 (-0.87,-0.55) | -0.97 (-1.07,-0.87) | -0.88 (-1.05,-0.70) |
| **Iceland** | -0.07 (-0.95,+0.90) | +0.05 (-0.34,+0.44) | +0.00 (-0.59,+0.58) | -0.20 (-1.18,+0.84) | -0.26 (-0.74,+0.19) | -0.22 (-0.90,+0.46) |
| **Italy** | -1.01 (-1.07,-0.95) | -1.05 (-1.33,-0.77) | -0.85 (-0.93,-0.78) | -1.25 (-1.31,-1.18) | -1.56 (-1.80,-1.34) | -1.36 (-1.45,-1.28) |
| **Lithuania** | -1.26 (-1.54,-0.97) | -1.21 (-1.36,-1.05) | NA | -1.69 (-2.00,-1.36) | -1.83 (-2.07,-1.59) | NA |
| **Netherlands** | -0.57 (-0.68,-0.46) | -0.66 (-0.76,-0.56) | -0.46 (-0.56,-0.36) | -0.80 (-0.92,-0.68) | -1.03 (-1.14,-0.92) | -0.89 (-1.00,-0.78) |
| **Norway** | +0.18 (-0.04,+0.40) | +0.07 (-0.01,+0.14) | -0.05 (-0.20,+0.10) | +0.18 (-0.04,+0.42) | +0.06 (+0.00,+0.13) | -0.26 (-0.44,-0.09) |
| **Poland** | -1.07 (-1.15,-1.00) | -1.02 (-1.16,-0.89) | -1.04 (-1.13,-0.95) | -1.50 (-1.58,-1.41) | -1.54 (-1.67,-1.41) | -1.57 (-1.68,-1.46) |
| **Portugal** | -0.69 (-0.84,-0.54) | -0.78 (-0.94,-0.61) | -0.60 (-0.72,-0.48) | -0.83 (-1.02,-0.67) | -0.93 (-1.03,-0.84) | -0.76 (-0.92,-0.60) |
| **Sweden** | -0.59 (-0.76,-0.46) | -0.54 (-0.63,-0.45) | -0.30 (-0.41,-0.20) | -0.87 (-1.02,-0.72) | -0.90 (-0.99,-0.81) | -0.72 (-0.83,-0.60) |
| **Slovenia** | -1.06 (-1.35,-0.71) | -1.02 (-1.18,-0.86) | -0.91 (-1.15,-0.68) | -0.94 (-1.29,-0.57) | -1.04 (-1.21,-0.88) | -1.08 (-1.35,-0.82) |
| **Slovakia** | -0.79 (-0.99,-0.58) | -0.85 (-1.00,-0.70) | -0.75 (-0.95,-0.54) | -0.83 (-1.07,-0.59) | -1.00 (-1.12,-0.88) | -0.91 (-1.14,-0.68) |
| **USA** | -1.65 (-1.68,-1.62) | -1.61 (-1.70,-1.51) | NA | -2.23 (-2.26,-2.20) | -2.27 (-2.39,-2.15) | NA |
| **Luxembourg** | NA | -0.55 (-0.84,-0.28) | -0.55 (-1.01,-0.08) | NA | -0.35 (-0.64,-0.07) | -0.69 (-1.21,-0.17) |
| **Ireland** | NA | NA | -0.36 (-0.54,-0.19) | NA | NA | -0.27 (-0.46,-0.08) |

**Figure A1.**
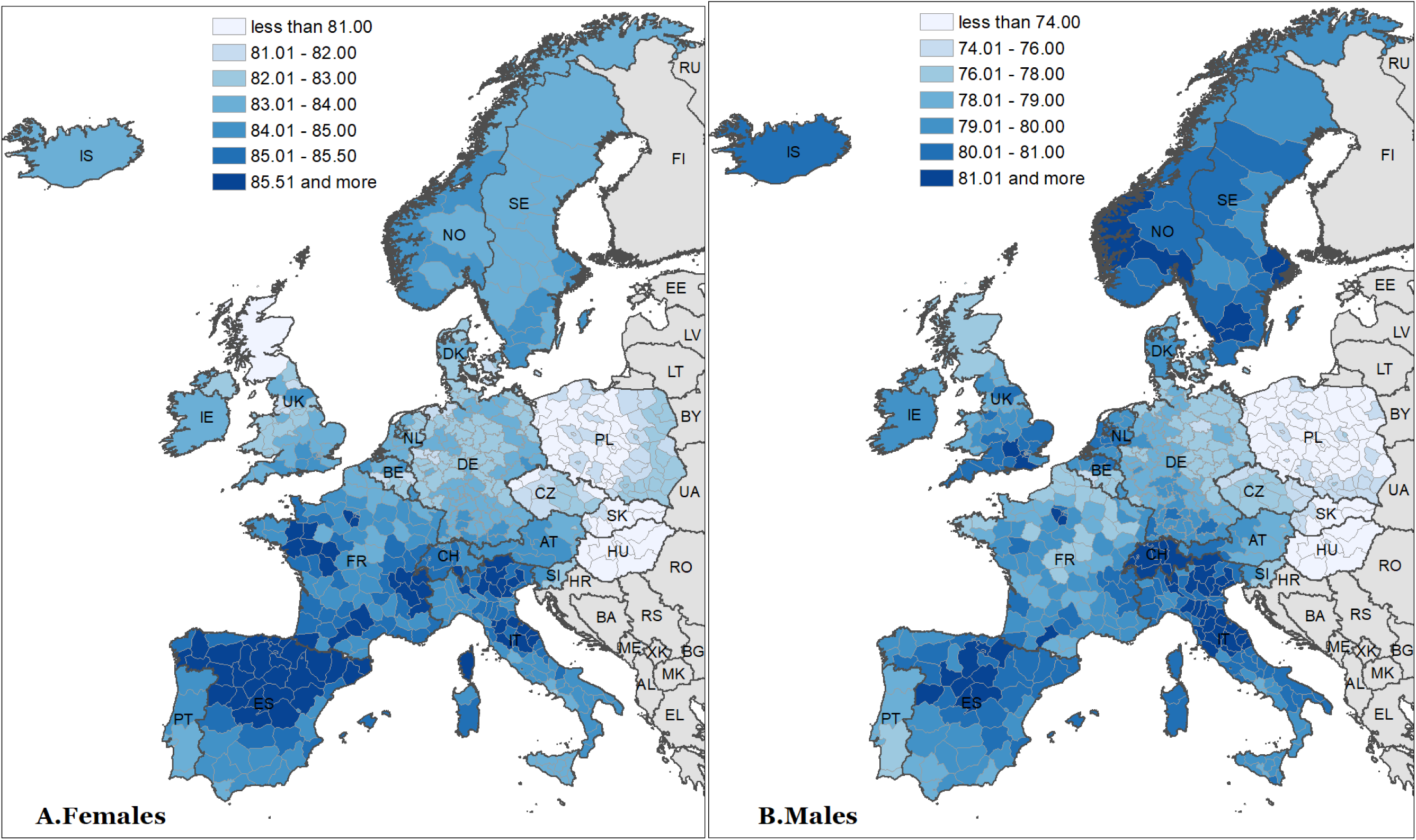
Life expectancy at birth across 561 spatial units in 21 European countries, 2015–2019.

**Figure A2.**
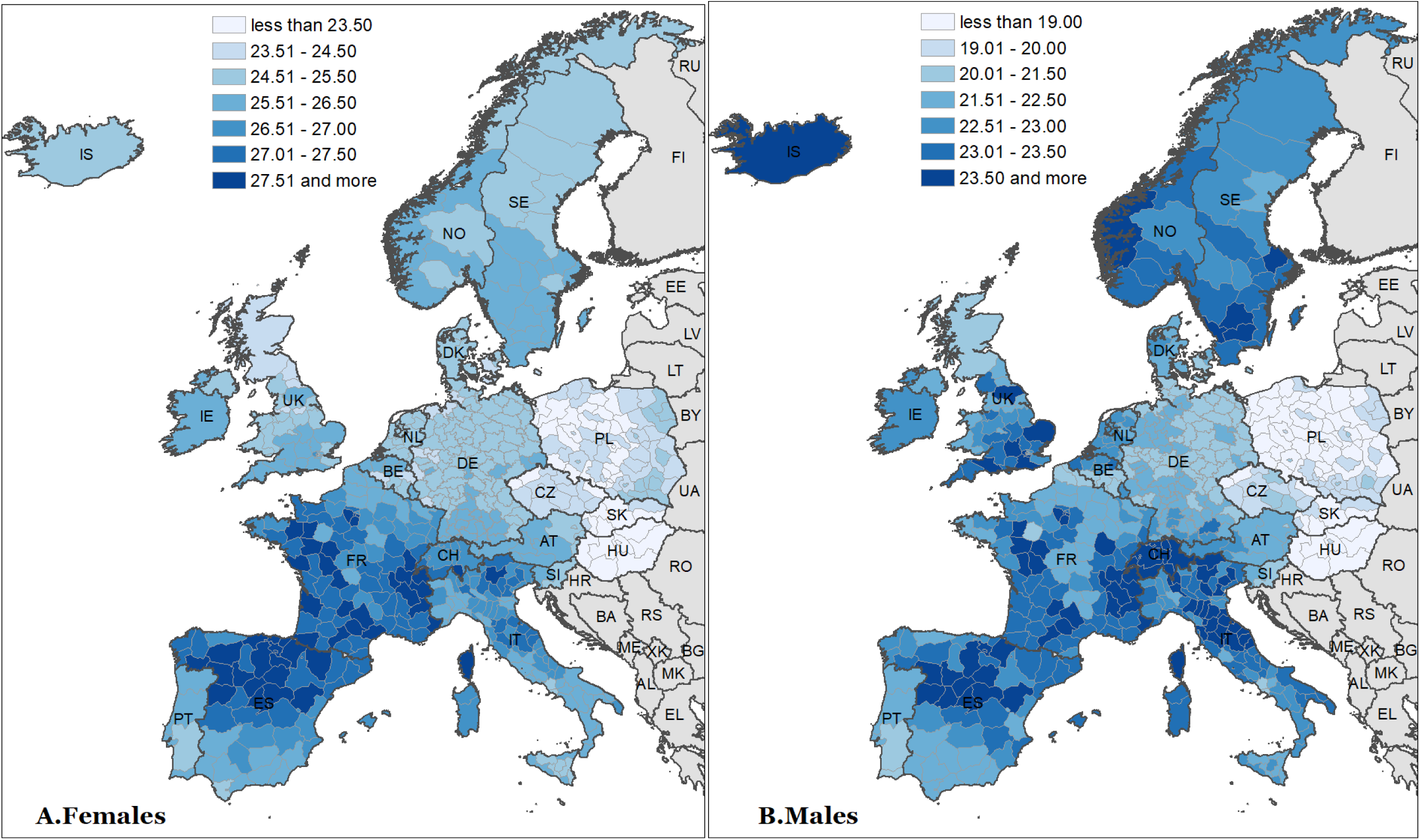
Life expectancy at age 60 across 561 spatial units in 21 European countries, 2015–2019.

**Figure A3.**
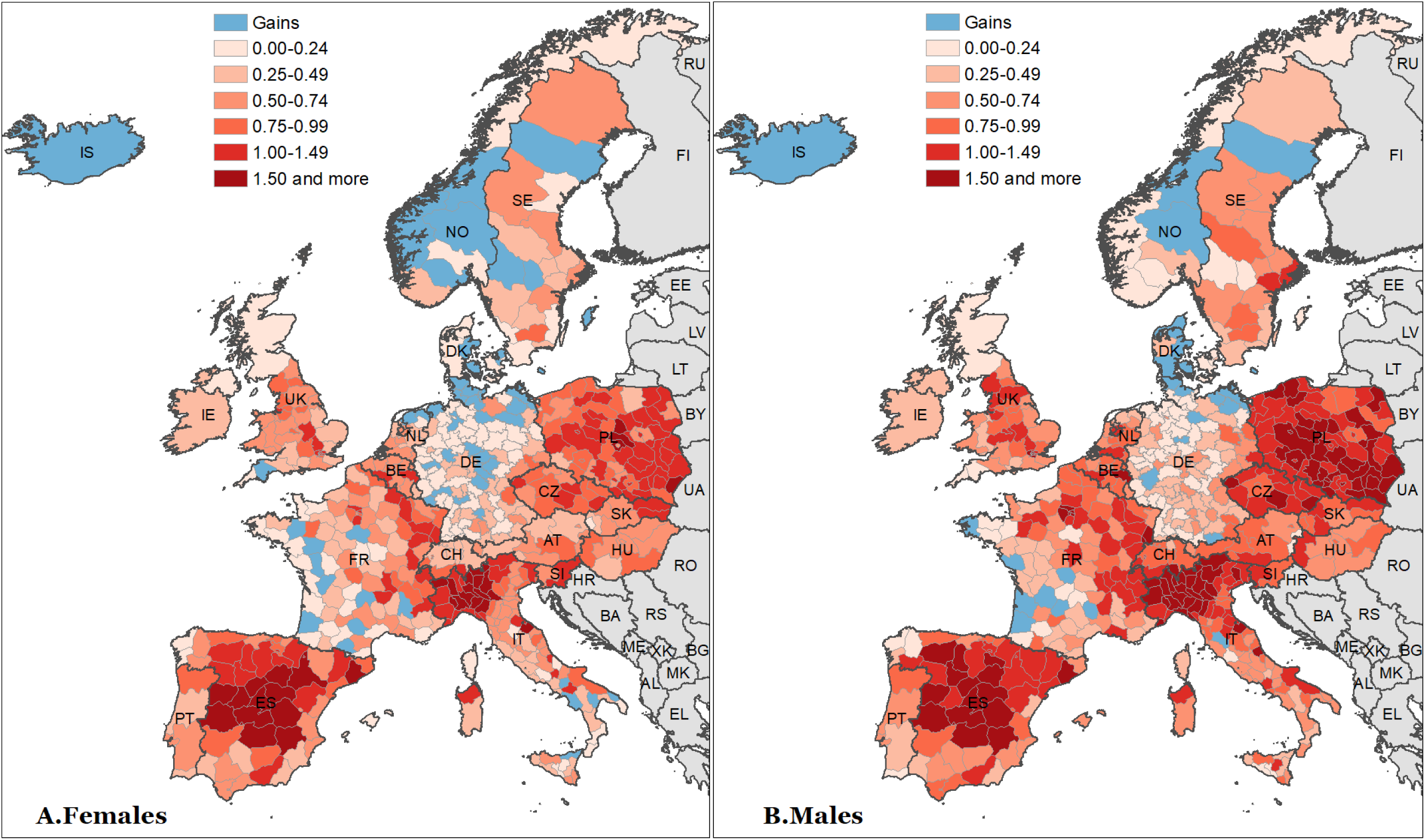
Spatial distribution of losses of life expectancy at age 60 across 21 European countries in 2020.

**Figure A4.**
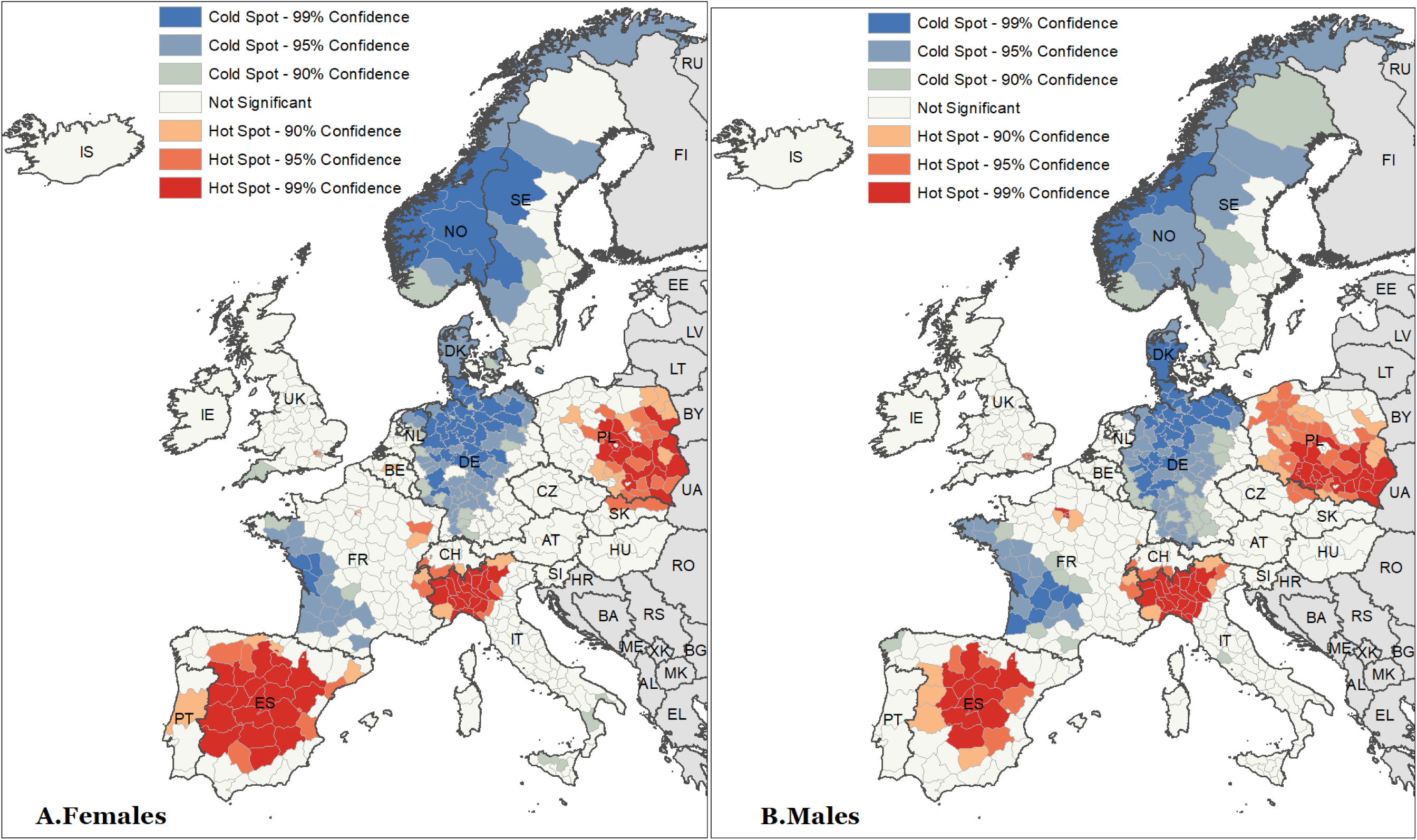
Hot and cold spots of losses of life expectancy at age 60 across 21 European countries in 2020.

**Figure A5.**
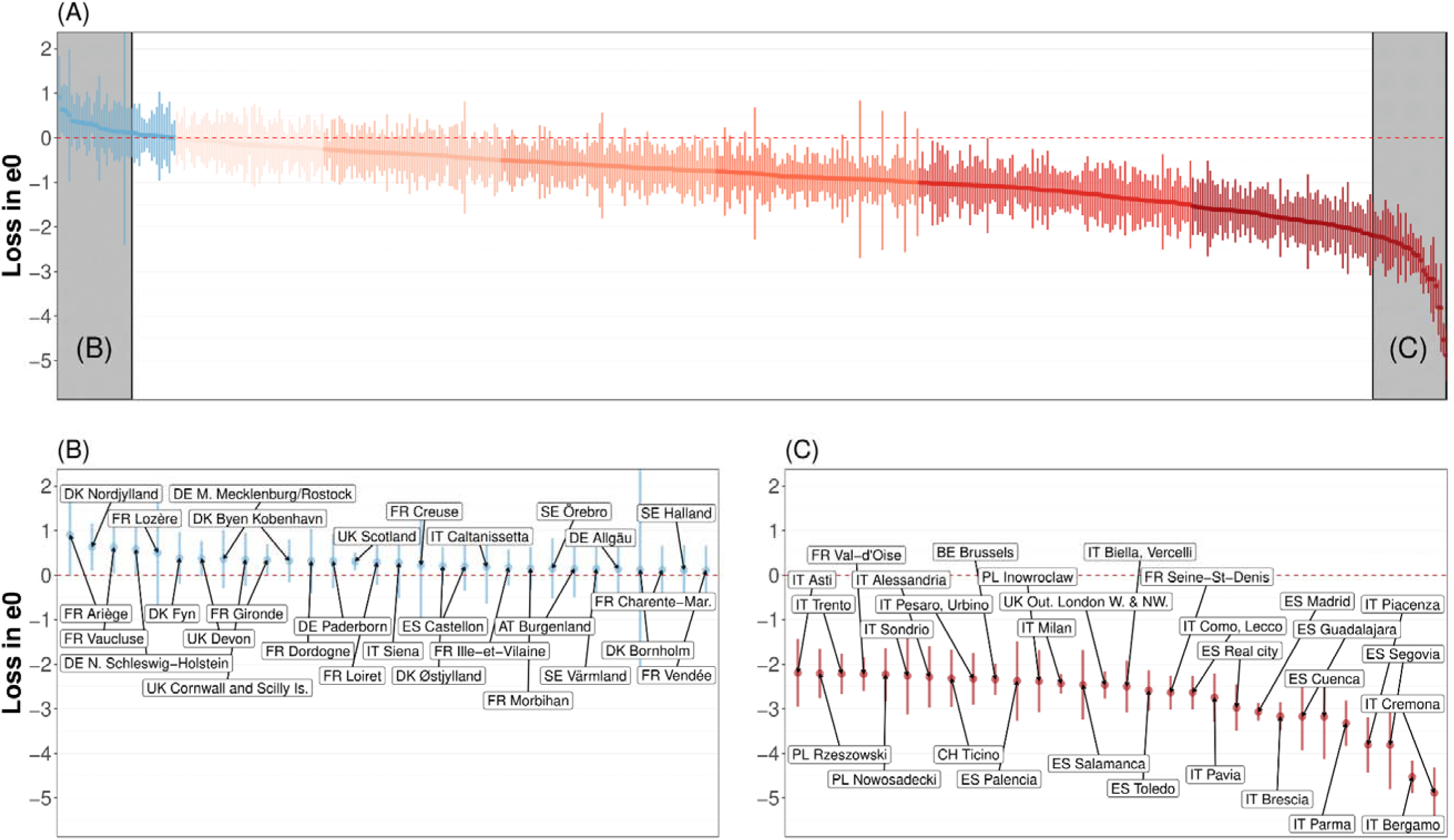
Losses or gains of life expectancy at birth (e0) associated with COVID-19 pandemic across 561 spatial units in 21 European countries, 2020, males.

**Figure A6.**
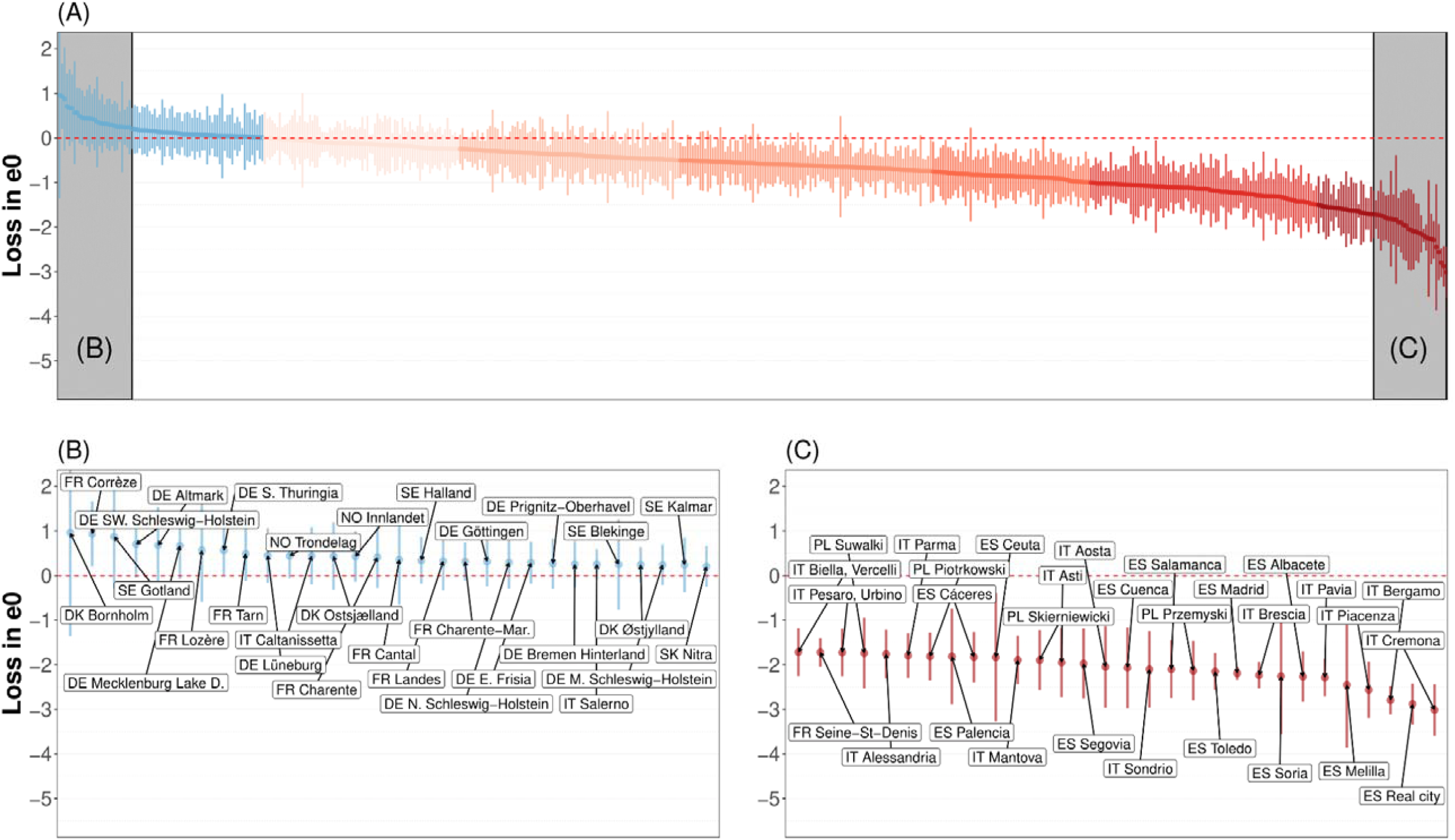
Losses or gains of life expectancy at birth (e0) associated with COVID-19 pandemic across 561 spatial units in 21 European countries, 2020, females.

## Online Supplementary Appendix B

Detailed values of our estimates and data visualisation tool are available at: https://osf.io/h68wz/?view_only=47353f6f4b2e41cab3c761246e59d615

Please read first “Online Appendix B.pdf”.

## Online Supplementary Appendix C

A detailed description of the analytic procedure to compute excess mortality is available at: https://osf.io/h68wz/?view_only=47353f6f4b2e41cab3c761246e59d615

## Notes

### Competing Interest Statement

The authors have declared no competing interest.

### Funding Statement

This study was funded from European Research Council (ERC) under the European Union Horizon 2020 research and innovation programme (grant agreement No 851485).

### Author Declarations

Data provided by national vital registration systems or Eurostat

### Summary of Updates

Change of Figure 3 Detailed methodology in Appendix Little changes in literature review

